# Leisure-time physical activity on lifelong trajectories of body mass index and obesity risk throughout life: multivariable regression and Mendelian randomization analyses using real-world data from the CORDELIA-Catalunya Study

**DOI:** 10.64898/2026.02.23.26346892

**Authors:** Javier Hernando-Redondo, Anna Camps-Vilaró, Roberto Elosua, Eleonora Fornara, Marcelino Bermúdez-López, Pere Torán⍰Monserrat, Ainara Jiménez-Navarro, José M. Valdivielso, Víctor M. López⍰Lifante, Tomàs Salas-Fernández, Serafí Cambray, Raquel Cruz, Jaume Marrugat, Álvaro Hernáez

## Abstract

**Background:** Evidence on how leisure-time physical activity (LTPA) improves lifetime body mass index (BMI) remains fragmented and prone to confounding.

**Methods:** We pooled 14,993 adults (30-90 y; 52.7% women; cohorts: REGICOR-ACRISC, ILERVAS, ARTPER) with baseline estimated LTPA (moderate-to-vigorous LTPA [MVLTPA] in REGICOR-ACRISC), genotype, and repeated BMI values from electronic health records (1990-2024, 36,157 measures). LTPA was categorized into cohort-specific quartiles; MVLTPA in 0, <100, <200, and ≥200 METs-min/day. In one-sample Mendelian randomization analyses, we categorized participants in quartiles of a cardiorespiratory fitness polygenic risk score derived from a large GWAS in UK Biobank. Group-dependent BMI trajectories were modeled using spline mixed-effects models. Obesity onset (first BMI ≥30 kg/m^2^) was analyzed with IPW-weighted Kaplan-Meier curves and Cox models.

**Results:** Higher LTPA was associated with slower BMI increases in ages 30-60 (Q1: +0.120 vs Q4: +0.075 kg/m^2^·year), slower declines in ages 70-90 (Q1: -0.143 vs Q4: -0.123 kg/m^2^·year), and lower obesity risk (Q4 vs Q1: HR 0.83, 95% CI 0.72-0.96). Similar trends were observed for MVLTPA. Higher genetically determined cardiorespiratory fitness showed parallel gradients (ages 30-60, Q1: +0.109 vs Q4: +0.101 kg/m^2^·year; ages 70-90, Q1: -0.130 vs Q4: -0.102 kg/m^2^·year) and lower obesity risk (Q4 vs Q1: HR 0.66, 0.56-0.78). Associations were present for women and men separately, but were stronger in men.

**Conclusions:** Higher LTPA and MVLTPA were associated with more favorable lifelong BMI trajectories, delayed obesity risk, and convergent support from Mendelian randomization analyses, supporting a causal protective role of physical activity (in both sexes but stronger in men).

## Introduction

High body mass index (BMI) is a major and growing global health concern, contributing substantially to morbidity, mortality, and disability across regions, sexes, and age groups [1,2]. Population-level prevention through healthy lifestyle habits is therefore a central public health priority [3]. Leisure-time physical activity (LTPA) is widely recommended for weight management, particularly moderate-to-vigorous LTPA (MVLTPA) [4]. However, the causal role of LTPA on BMI has only been shown in short- or medium-term randomized controlled trials focusing on specific age groups [5–7]. The only evidence covering longer follow-up periods is observational, which is susceptible to residual and unmeasured confounding and does not demonstrate causality (individuals meeting LTPA recommendations often have healthier diets, smoke less, have higher socioeconomic status, and better underlying health), and has not covered the entire life course [8,9]. In the absence of long-term randomized trials specifically addressing lifelong effects of LTPA on adiposity, triangulation of observational evidence with other robust causal inference tools becomes essential [10]. In this context, real-world evidence derived from electronic health records may serve beneficial to this purpose, as it has been previously used to characterize the worldwide epidemiologic landscape of overweight/obesity [11].

Mendelian randomization (MR) uses genetic variants as instruments to estimate the unconfounded effects of modifiable exposures on outcomes [12]. When multivariable regression and MR results agree, this increases confidence that the association is causal [10]. Yet, applying MR directly to self-reported LTPA is challenging because physical activity is a complex behavioral trait that is not likely to be explained by genetic predisposition, raising concerns about instrument strength and MR assumptions [13]. Nevertheless, we could overcome this limitation by using physiological proxies that are tightly linked to habitual LTPA and have stronger genetic architecture, such as cardiorespiratory fitness (CRF) [14]. High CRF reflects the integrated response of cardiovascular and muscular systems to habitual exercise [15], can increase with higher LTPA as well as due to genetic predisposition, and is therefore suitable for MR analyses under our hypothesis. Thus, in this study we aimed to assess whether self-reported LTPA and MVLTPA levels and genetically determined CRF are associated with differences in lifelong BMI trajectories and the risk of incident obesity in the CORDELIA-Catalunya Study (in all participants, and in women and men separately).

## MATERIALS AND METHODS

### Study design and population

We conducted a prospective observational study using data from the three existing Catalan cohorts with self-reported LTPA information, genotype data, and repeated measures of BMI included in the CORDELIA project: REGICOR-ACRISC, ILERVAS, and ARTPER [16]. Eligible participants had a baseline estimate of self-reported LTPA record, genotype data, and at least two measures of BMI over time (**Figure 1**).

**Figure 1.**
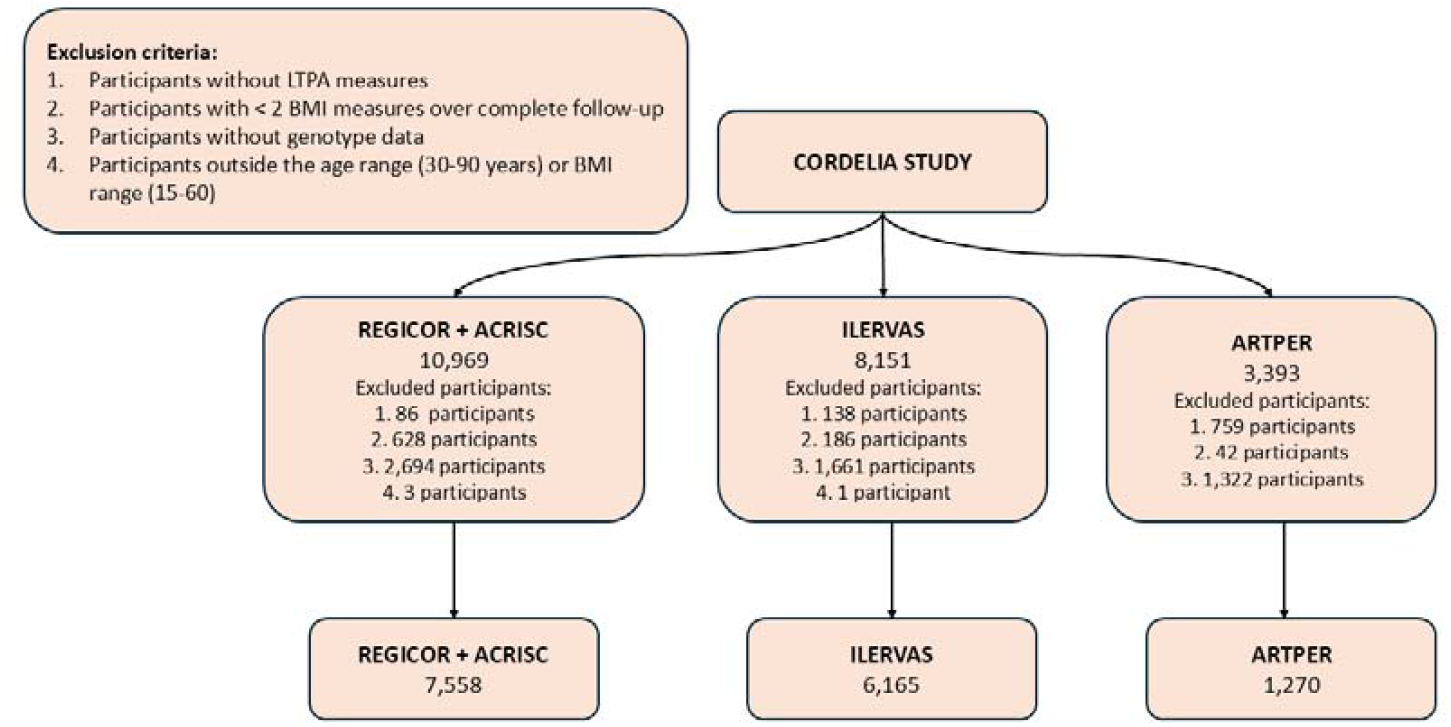
Flow chart.

### Leisure-time physical activity

LTPA was estimated by the long version of Minnesota questionnaire in REGICOR-ACRISC [17,18], the short version of Minnesota questionnaire in ARTPER [19], and the long version of the IPAQ questionnaire in ILERVAS [20]. The questionnaires registered the frequency and duration of 67 (Minnesota, long version), 27 (IPAQ, long version), and 6 activities (Minnesota, short version) undertaken by the participants during the previous year. LTPA was expressed as metabolic equivalents of task-minute per day (MET-min/day), calculated by multiplying the METs linked to each activity by its average minutes per day and summing across all activities. METs-min/day due to moderate and vigorous activities were used to estimate MVLTPA levels in REGICOR.

### Genetic predisposition for cardiorespiratory fitness

We used the results from the most recent genome-wide association study (GWAS) of objectively measured CRF that presented sex-stratified results to create the genetic instruments for our MR analyses [14]. This GWAS included 70,783 participants, aged 40-69, living in the United Kingdom (none of whom participated in the CORDELIA Study), with available genotype data and CRF measured as maximal oxygen consumption during a ramp test [14]. It identified genetic variants linked to CRF in all participants and in women and men separately. We computed participants’ polygenic risk scores (PRS) for CRF using PRS-CS, which fits a Bayesian continuous-shrinkage model to GWAS summary statistics while incorporating an external linkage disequilibrium reference to jointly estimate the effect sizes of the genetic variants on CRF [21]. Each individual’s PRS was calculated as the allele dosage-weighted sum of these effects and was directly proportional to the genetically determined CRF levels.

### Body mass index and first occurrence of obesity

BMI data was obtained from the Public Data Analysis for Health Research and Innovation Program of Catalonia (PADRIS) at the Agency for Health Quality and Assessment of Catalonia. PADRIS is focused in real-world data exploitation, and routinely links individual-level demographic, socioeconomic, and healthcare data (including primary care and hospital visits) generated from consultations within the Catalan healthcare system [22]. All data were anonymized, de-identified, and accessed in accordance with applicable regulations, ethical guidelines, and transparency principles. BMI (in kg/m^2^) was calculated for each participant from PADRIS height and weight values. We excluded specific BMI measurements that differed by more than 12 kg/m^2^ from both the immediately preceding and the subsequent visit. Repeated BMI measures from 1990 to 2024 within the range 15-60 kg/m^2^ were included. Each participant contributed repeated BMI measurements only within the ages captured during their individual follow-up. By stitching together these partially overlapping age segments across all participants, we assembled a longitudinal dataset that collectively spans ages 30 to 90 years. We restricted the analyses to individuals aged 30-90 due to limited amount of BMI measures for participants aged 20-29 (1.1%) and 90-100 (0.03%).

The number of BMI observations available for each age is reported in **Supplementary Table 1**. From these data, we defined the first occurrence of obesity as the first recorded BMI >30 kg/m^2^ in participants without the condition at baseline.

### Other variables

The following information has been harmonized for all the participants of the participating cohorts in the beginning of their follow-up: age (continuous), prevalence of diabetes (yes/no), prevalence of hypertension (yes/no), prevalence of hypercholesterolemia (yes/no), smoking status (never smoker, current smoker, and former smoker [no smoking in the last 12 months]), and a proxy for socioeconomic status (area-level income indicator: median of the municipality-specific annual mean net per-capita income for the participant’s municipality of residence, 2015-2023) [23]. The first 10 ancestry-informative genetic principal components (PCs) were also gathered to account for population stratification in MR analyses.

### Ethical aspects

This study was performed in line with the principles of the Declaration of Helsinki. The study protocol was approved by the Ethics Committee of the Parc de Salut Mar (reference 2024/11670/I; date: November 18, 2024). Approval for the CORDELIA Study was granted by the Ethics Committee of the Parc de Salut Mar (2023/10785/I, date: March 29, 2023). All cohorts obtained informed consent from their participants.

### Statistical analysis

We described normally distributed continuous variables with means ± standard deviations (SD), and categorical variables with proportions.

#### Multivariable regression

We modeled BMI trajectories in the longitudinal, aggregated dataset spanning ages 30-90 using smoothed cubic spline mixed-effects regression models, with participants as random effects [24,25]. In analyses with LTPA as the exposure, we modeled BMI trajectories across four cohort-specific quartiles of LTPA (to account for between-cohort differences in the distribution and measurement of LTPA) [26]. In analyses with MVLTPA, we modeled four categories (0 METs-min/d [null MVLTPA, reference], <100 METs-min/d [insufficient MVLTPA], 100-<200 METs-min/d [adequate MVLTPA], and ≥200 METs-min/d [above-adequate]) among REGICOR-ACRISC participants. To allow different trajectories, models incorporated an interaction between age (the underlying time scale) and the LTPA/MVLTPA group. Analyses were adjusted for birth year, sex, and baseline prevalent diabetes, hypertension, hypercholesterolemia, smoking status, and socioeconomic status. Before the analyses, we imputed missing data using a nonparametric random forest-based approach in the following covariates: diabetes (0.16%), hypercholesterolemia (0.28%), hypertension (0.09%) and smoking status (0.56%). We graphed mean BMI trajectories by group using model-based predictions and estimated average between-group differences (with the lowest LTPA/MVLTPA group as the reference) at benchmark ages of 30, 40, 50, 60, 70, 80 and 90 via linear regressions. As a sensitivity analysis, we assessed whether sex modified the associations by fitting models that added a “sex × LTPA/MVLTPA group” interaction and compared them with the base models using likelihood ratio tests.

We also assessed the association between LTPA/MVLTPA and the incidence of obesity (first BMI ≥30 kg/m^2^). Using the same LTPA quartiles and MVLTPA categories defined for the trajectory analyses, we plotted cumulative incidence from ages 30-90 years for LTPA/MVLTPA groups using Kaplan-Meier curves. To reduce confounding in these curves, we applied inverse probability weighting based on a propensity score that included the same covariates as in the trajectory models, and we displayed weighted survival estimates with 95% confidence bands. Additionally, we estimated hazard ratios for obesity for LTPA/MVLTPA groups (relative to the lowest LTPA/MVLTPA group) using Cox proportional hazards models adjusted for the same covariates. As sensitivity analyses, we also investigated effects stratifying by sex.

#### Mendelian randomization analyses

We repeated the trajectory and incidence analyses using a one-sample MR framework, classifying participants into four groups according to quartiles (1-4) of the CRF-PRSs as the exposure in both analyses (trajectories: smoothed cubic spline mixed-effects models; incidence of obesity: weighted Kaplan-Meier curves and Cox proportional hazards models). Both analyses were adjusted only for the first 10 ancestry-informative genetic PCs.

We verified the three key MR assumptions in our analyses: i. the genetic instrument is robustly associated with the exposure (relevance); ii. there is no confounding of the instrument-outcome association by population stratification; and iii. the instrument affects the outcome only through the exposure (lack of horizontal pleiotropy) [13]. Regarding the relevance assumption, we quantified the strength of association between the CRF-PRS and CRF levels (estimated according to age, sex, BMI, heart rate, and MVLTPA [15]) reporting the F-statistic and the magnitude of the association [13]. We minimized confounding of the genetic instrument-outcome association due to population stratification by adjusting all MR analyses for the first 10 ancestry-informative PCs [27]. We assessed the lack of horizontal pleiotropy using several checks [28–32]. First, we evaluated the lack of association between the exposure in MR analyses (CRF-PRS quartiles) and the potential sources of horizontal pleiotropy, including baseline covariates in multivariable regression (sex, prevalent diabetes, prevalent hypertension, prevalent hypercholesterolemia, smoking status, and socioeconomic status), as well as age, using linear regressions and controlling for multiple comparisons with the Bonferroni method. Second, to examine whether any association between the CRF-PRS and BMI operated independently the estimated CRF and MVLTPA, we fitted a model for the cumulative average BMI (mean of all available BMI measures per participant, as the outcome) that included CRF-PRS quartiles (main exposure) and estimated CRF quartiles (additional covariate). Attenuation toward the null supported lack of horizontal pleiotropy. Finally, as a negative-control analysis, we verified the lack of association between the CRF-PRS and a variable theoretically unrelated to genetics (birth year), as an indication of no residual structure or broad pleiotropy.

All analyses were performed using R version 4.4.2. Code for data management and analysis is available at https://github.com/XXX.

## RESULTS

### Participants

Our population was 14,993 participants. The mean age at baseline was 53.5 years, and 52.7% were women. Characteristics of the participants at baseline of their respecting follow-ups is presented in **Table 1** and stratified by sex and cohort in **Supplementary Tables 1** and **2**, respectively. The median number of BMI records per individual was 7 in REGICOR (interquartile range = 7), 4 in ACRISC (interquartile range = 5), 7 in ILERVAS (interquartile range = 5), and 10 in ARTPER (interquartile range = 8). Trajectory analyses were based on 36,157 repeated measures (4.6% for ages 30-39, 15.7% for ages 40-49, 28.0% for ages 50-59, 27.0% for ages 60-69, 17.5% for ages 70-79, and 6.9% for ages 80-89).

**Table 1.** Main cardiovascular risk factors.

| Variable | Value |
| --- | --- |
| Age at baseline, years (mean ± SD) | 53.5 ± 11.0 |
| Female sex (*n*, %) | 7,903 (52.7%) |
| Baseline BMI, kg/m<sup>2</sup> (mean ± SD) | 27.5 ± 4.61 |
| Baseline BMI categories: |  |
| Normal weight (<25 kg/m<sup>2</sup>) (*n*, %) | 4,616 (30.8%) |
| Overweight (25.0-29.9 kg/m<sup>2</sup>) (*n*, %) | 6,512 (43.4%) |
| Obesity (≥30 kg/m<sup>2</sup>) (*n*, %) | 3,865 (25.8%) |
| Baseline prevalence of diabetes (*n*, %) | 1,000 (6.67%) |
| Baseline prevalence of hypertension (*n*, %) | 5,342 (35.6%) |
| Baseline prevalence of hypercholesterolemia (*n*, %) | 6,269 (41.8%) |
| Smoking status: |  |
| Current smoker (*n*, %) | 3,853 (25.7%) |
| Never smoked (*n*, %) | 7,134 (47.6%) |
| Former smoker (*n*, %) | 4,006 (26.7%) |
| Socioeconomic status, €/year (median, 1<sup>st</sup>-3<sup>rd</sup> quartile) | 13,161 (11,949; 14,910) |

### Lifelong BMI trajectories and risk of obesity according to LTPA and MVLTPA groups

Participants in the groups with higher LTPA tended to increase BMI more slowly between ages 30-60 (Q1: +0.12 kg/m^2^ × year, 95% CI 0.118 to 0.121; Q2: +0.122 kg/m^2^ × year, 95% CI 0.121 to 0.124; Q3: +0.099 kg/m^2^ × year, 95% CI 0.097 to 0.101; Q4: +0.075 kg/m^2^ × year, 95% CI 0.073 to 0.077). Conversely, participants in the groups with higher LTPA tended to show a slower decline in BMI over time between ages 70-90, particularly among those above the median LTPA (Q1: -0.143 kg/m^2^ × year, 95% CI -0.145 to -0.140; Q2: -0.125 kg/m^2^ × year, 95% CI -0.128 to -0.122; Q3: -0.108 kg/m^2^ × year, 95% CI -0.111 to -0.105; Q4: -0.123 kg/m^2^ × year, 95% - 0.126 to -0.120 CI) (**Figure 2A**). This translated into lower predicted BMI values in the groups with higher LTPA between ages 50-90 (**Figure 2B**). These differences also translated into lower incidence of first occurrence of obesity relative to Q1 (median time to event 66.9 years) in Q2 (HR 0.77, 95% CI 0.67 to 0.90; median time to event 72.5 years), Q3 (HR 0.82, 95% CI 0.71 to 0.95; median time to event 73.4 years), and Q4 (HR 0.83, 95% CI 0.72 to 0.96; median time to event 70.1 years) (**Figure 2C**).

**Figure 2.**
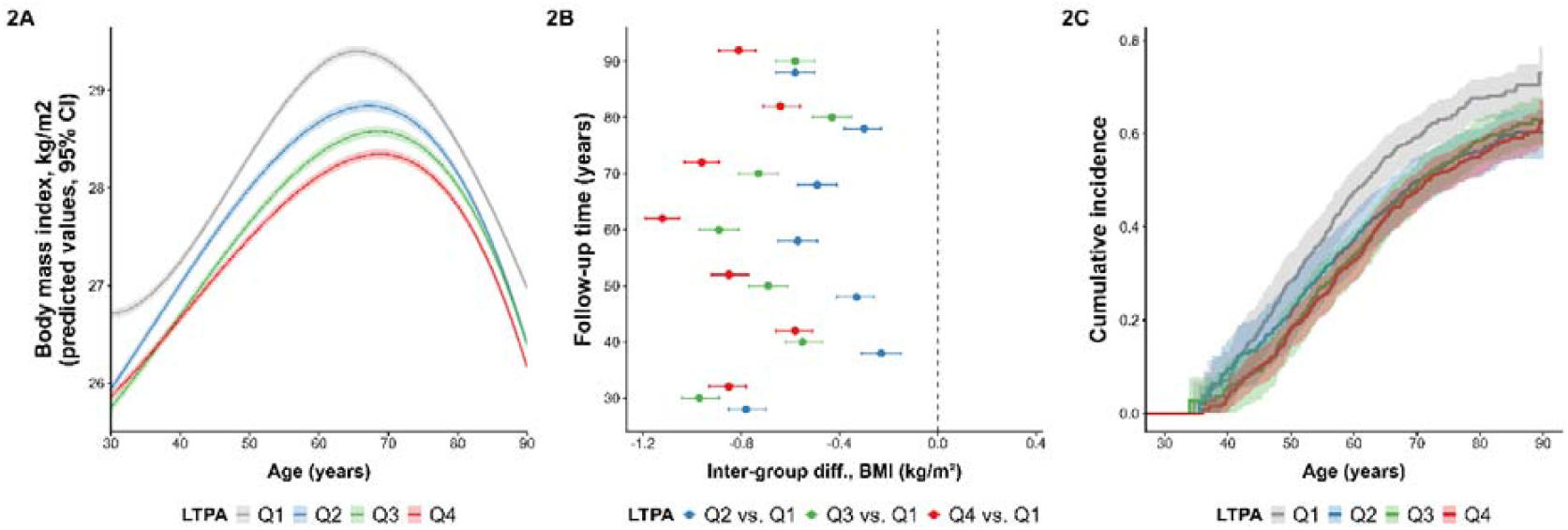
BMI trajectories and incident obesity by cohort-specific quartiles of LTPA (Q1 in grey, Q2 in blue, Q3 in green, Q4 in red). A. BMI trajectories per quartiles. B. Inter-group differences in predicted mean BMI values between ages 30-90. C. Weighted Kaplan-Meier curves for first obesity onset.

Participants in groups with higher MVLTPA tended to increase BMI more slowly between ages 30-60, particularly at MVLTPA ≥100 METs-min/day (0 METs-min/day: +0.116 kg/m^2^ × year, 95% CI 0.114 to 0.118; >0 to <100 METs-min/day: +0.124 kg/m^2^ × year, 95% CI 0.122 to 0.126; ≥100 to <200 METs-min/day: +0.085 kg/m^2^ × year, 95% CI 0.083 to 0.088; ≥200 METs-min/day: +0.087 kg/m^2^ × year, 95% CI 0.085 to 0.089) (**Figure 3A**). This translated into lower predicted BMI values in the groups with adequate and above-adequate MVLTPA between ages 50-90 (**Figure 3B**). Kaplan-Meier curves also suggested adequate MVLTPA levels were also linked to lower incidence of first occurrence of obesity relative to null MVLTPA (median time to event 65.41 years) in individuals with >0 to <100 METs-min/day (insufficient MVLTPA, HR 0.73, 95% CI 0.63 to 0.85; median time to event 71.8 years), ≥100 to <200 METs-min/day (adequate MVLTPA, HR 0.62, 95% CI 0.53 to 0.74; median time to event 81.6 years), and ≥200 METs-min/day (above-adequate MVLTPA, HR 0.66, 95% CI 0.56 to 0.78; median time to event 71.5 years) (**Figure 3C**).

**Figure 3.**
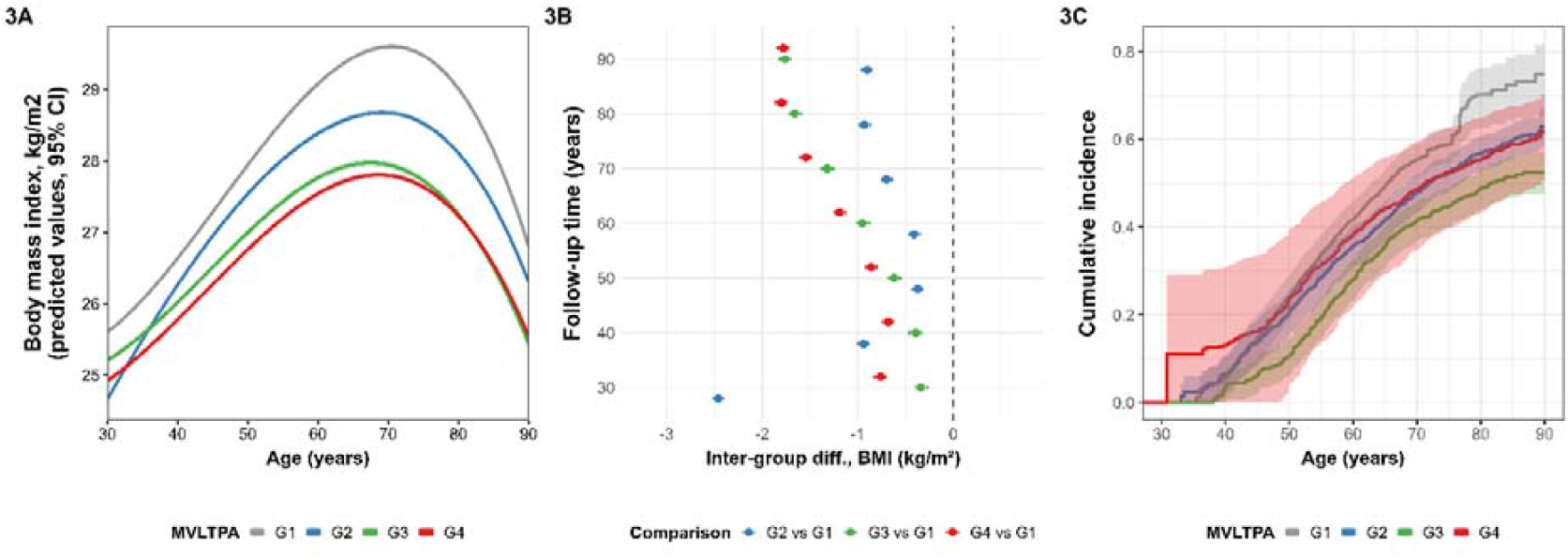
BMI trajectories and incident obesity by MVLTPA levels in REGICOR-ACRISC (0 METs-min/day in grey, >0 to <100 METs-min/day in blue, ≥100 to <200 METs-min/day in green, ≥200 METs-min/day in red). A. BMI trajectories per quartiles. B. Inter-group differences in predicted mean BMI values between ages 30-90. C. Weighted Kaplan-Meier curves for first obesity onset.

### Genetically determined cardiorespiratory fitness, BMI trajectories, and risk of obesity

Participants with a genetic predisposition to higher CRF tended to show slower increases in BMI between ages 30 and 60 (Q1: +0.109 kg/m^2^ × year, 95% CI 0.109 to 0.110; Q2: +0.108 kg/m^2^ × year, 95% CI 0.107 to 0.109; Q3: +0.094 kg/m^2^ × year, 95% CI 0.093 to 0.095; Q4: +0.101 kg/m^2^ × year, 95% CI 0.100 to 0.102). In addition, participants with the highest genetically determined CRF showed an attenuated decrease in BMI over time between ages 70 and 90 (Q1: -0.130 kg/m^2^ × year, 95% CI -0.131 to -0.129; Q2: -0.119 kg/m^2^ × year, 95% CI - 0.120 to -0.117; Q3: -0.144 kg/m^2^ × year, 95% CI -0.146 to -0.142; Q4: -0.102 kg/m^2^ × year, 95% CI -0.103 to -0.100) (**Figure 4A**). Thus, genetic predisposition to higher CRF was associated with lower predicted BMI among groups over time (**Figure 4B**). These differences were also associated with a lower incidence of first-onset obesity relative to Q1 (median time-to-event 65.0 years) in Q2 (HR 0.75, 95% CI 0.64 to 0.87; median time-to-event 72.5 years), Q3 (HR 0.67, 95% CI 0.56 to 0.80; median time-to-event 79.0 years), and Q4 (HR 0.66, 95% CI 0.56 to 0.78; median time-to-event 76.7 years) (**Figure 4C**).

**Figure 4.**
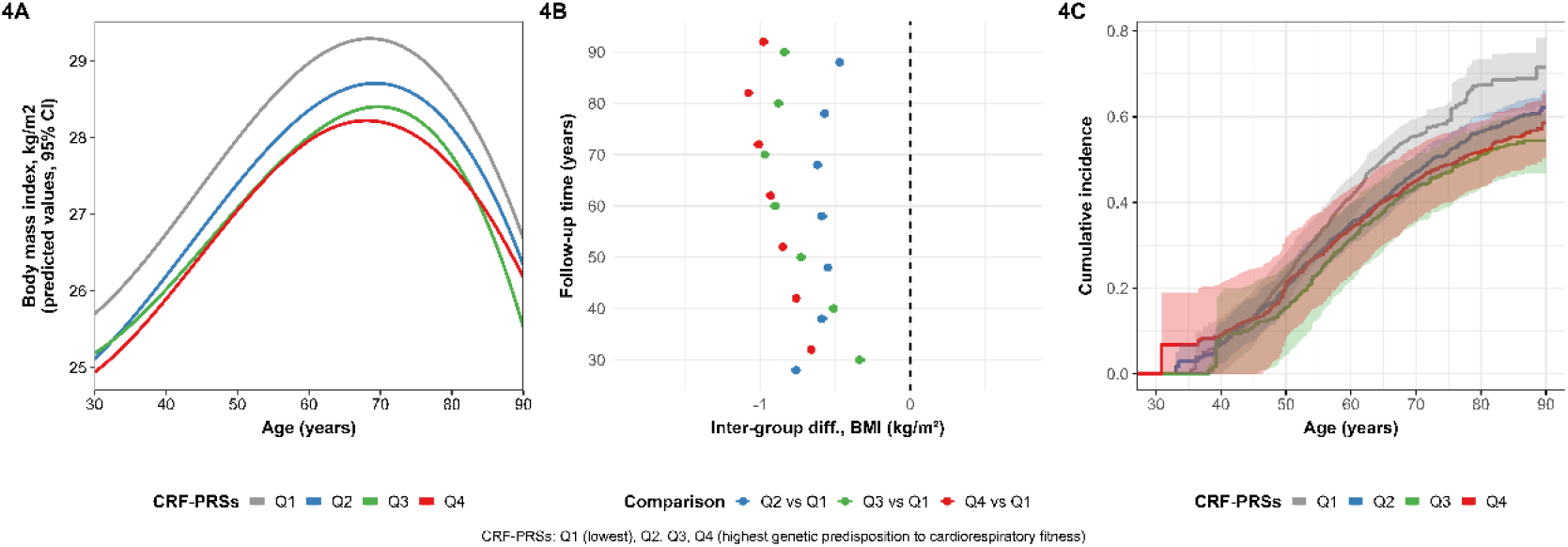
BMI trajectories and incident obesity by genetically determined CRF levels (Q1 in grey, Q2 in blue, Q3 in green, Q4 in red). A. BMI trajectories per quartiles. B. Inter-group differences in predicted mean BMI values between ages 30-90. C. Weighted Kaplan-Meier curves for first obesity onset.

MR results supported the validity of the analyses, with no evidence of violations of the key assumptions. The genetic instrument (CRF-PRS) was associated with the phenotype it represents (estimated CRF): F-statistic = 7.66; β = 0.32, 95% CI 0.094 to 0.55), and the adjustment for the first 10 genetic PCs was used to minimize the bias due to population structure in our analyses. Regarding horizontal pleiotropy, no meaningful associations were observed between CRF-PRS quartiles and the pre-identified sources of pleiotropy, or in the negative-control analysis (birth year) (*p*-values were > 0.00625, considering the Bonferroni method for eight comparisons): age (*p*-value = 0.850), sex (*p*-value = 0.222), diabetes (*p*-value = 0.217), hypertension (*p*-value = 0.645), hypercholesterolemia (*p*-value = 0.641), socioeconomic status (*p*-value = 0.093), smoking (*p*-value = 0.009), and birth year (*p*-value = 0.619). In addition, adding CRF quartiles to the PRS-cumulative BMI mean showed the association’s attenuation toward the null (unadjusted β = -0.28, 95% CI -0.36 to -0.20; adjusted β = -0.17, 95% CI -0.26 to -0.076; attenuation: 40%).

### Sex-specific analyses

LTPA analyses in women and men separately also showed a progressive attenuation in the BMI increase between ages 30-60, as well as a decreased risk of obesity in higher LTPA groups, with a stronger and more graded association in men (only women, Q4 vs Q1: HR 0.85, 95% CI 0.73 to 0.99; only men, Q4 vs Q1: HR 0.75, 95% CI 0.65 to 0.87) (**Supplementary Figures 1 and 2**). MVLTPA analyses in women and men separately showed similar findings, as observed in obesity incidence analyses, again with a stronger association in men (only women, adequate vs. no MVLTPA: HR 0.64, 95% CI 0.51 to 0.80; above-adequate vs. no MVLTPA: HR 0.70, 95% CI 0.54 to 0.89; only men, adequate vs. no MVLTPA: HR 0.58, 95% CI 0.45 to 0.75; above-adequate vs. no MVLTPA: HR 0.60, 95% CI 0.47 to 0.76;) (**Supplementary Figures 3 and 4**). Sex-specific MR results also point in the same direction, with a lower risk of obesity in individuals in individuals with higher genetically determined CRF that is significant in both women and men separately but of greater magnitude among men (only women, Q4 vs Q1: HR 0.73, 95% CI 0.56 to 0.94; only men, Q4 vs Q1: HR 0.61, 95% CI 0.48 to 0.77) (**Supplementary Figures 5 and 6**).

## DISCUSSION

By integrating LTPA and MVLTPA data, genotype information, and long-term repeated BMI measurements, we were able to estimate lifelong trajectories and strengthen the evidence for a causal, protective role of physical activity against obesity over the lifespan. Across complementary analyses, higher LTPA and MVLTPA were consistently associated with a slower increase from early-to-mid adulthood, slower BMI decreases from in older adults, lower predicted BMI from midlife onwards and a delayed and lower risk of first onset obesity in general population. The concordant direction and shape of associations across multivariable regression models and one-sample Mendelian randomization support the interpretation that physical activity causes a more favorable BMI evolution over time. These patterns were observed in women and men, with overall stronger associations in men.

Our findings align with prior observational work indicating that higher LTPA is linked to lower adiposity and healthier weight profiles over time [33], that physical activity is also associated with a slower BMI decline in aging populations [34], and agree with longitudinal evidence considering more than baseline LTPA values [35]. Taken together with complementary MR evidence, our data support a causal role of LTPA/MVLTPA and its consequences (fitness) in shaping long-term BMI evolution and obesity risk. We extend current evidence of short-to medium-term effects of exercise training on reduced adiposity and improved weight-related phenotypes to longer term follow-up [5–7] supporting causality across the life course. MR studies of activity-related traits and objective fitness indicators generally support a causal role in lifelong lower BMI and fat body mass [Fornara E, Med Sci Sport Exerc, 2026, in press]. Besides previous evidence in twin studies pointing to a causal role of physical activity on lower BMI [36], our one-sample MR using genetically determined CRF adds to this causal triangulation: CRF is a plausible physiological proxy for habitual exercise exposure with stronger genetic architecture than self-reported activity [14], and the concordance between CRF-PRS gradients and both BMI trajectories and obesity incidence supports biological plausibility. There are several mechanistic evidence that also support this causal role, as higher activity/fitness: (i) increases total daily energy expenditure [37]; (ii) promotes maintenance and increases of lean muscle mass [38], which can contribute to higher basal energy expenditure over time and could also help explain the slower BMI decline in older populations due to sarcopenia; and (iii) activates AMP-mediated protein kinase, improving fatty acid oxidation and mitochondrial function in ways that limit long-term adiposity [39].

The moderately stronger associations we observed in men (together with beneficial associations in both women and men separately) are compatible with prior literature. Some large cohort analyses have reported slightly stronger associations in men between physical activity expenditure and other cardiometabolic health outcomes such as type-2 diabetes [40], whereas exercise trials suggest that, when prescribed energy expenditure is comparable, sex differences in body-weight change are often small [41]. Biologically, larger BMI separation in men in free-living settings could reflect (i) higher lean mass, so the same reported MET-min/day implies higher absolute energy expenditure; (ii) sex differences in fat distribution/mobilization, with men tending to mobilize more visceral fat during negative energy balance [42]; and (iii) female life-course BMI evolution and menopause-related shifts in body composition that may moderate BMI-based differences in women despite similar metabolic benefits [42,43]. Methodologically, self-reported LTPA/MVLTPA may be differentially misestimated by sex [44], BMI may not fully reflect beneficial changes in body composition (e.g., fat loss with preserved or increased muscle), and the same questionnaire-based activity may correspond to different energy expenditure in men and women [45]. Strengths of our study include the ability to integrate repeated real-world BMI measures across three decades, sex-stratified analyses, and, critically, the triangulation of observational and genetic-proxy evidence within the same framework to suggest that residual confounding is not likely to explain the physical activity-associated benefits on BMI trajectories and obesity risk over time. However, our work also has several limitations. First, BMI trajectories were assembled by stitching partially overlapping age segments contributed by different individuals and time periods, creating potential temporal heterogeneity that could influence the reconstructed “population trajectory”. Because each participant contributed data only during the age windows in which they had primary care encounters, rather than being observed continuously across the entire 30-90 year span, our approach implicitly assumes that missing BMI values are missing at random and that censoring/loss to follow-up is minimized after covariate adjustment. To mitigate potential bias, our spline mixed-effects models used all available repeated BMI measures without requiring every individual to contribute data at every age, analyses were restricted to individuals that contributed with at least two valid measures of BMI over time, age-as-time-scale survival models treat loss to follow-up as right censoring, adjustment for birth year aimed to minimize time heterogeneity, and we rebalanced obesity incidence curves using inverse probability weighting based on measured predictors. Nevertheless, prospective studies with more complete long-term follow-up and minimal attrition are needed to confirm these reconstructed life-course patterns. Second, BMI in routine care is measured at clinically driven encounters; if visit frequency depends on underlying weight change or health deterioration, it can imply an overrepresentation of individuals with more unfavorable BMI values and can bias trajectory estimation. Third, selection and survivor biases are possible in late-life follow-up (healthier individuals contribute more measurements at older ages), and obesity onset based on first recorded BMI ≥30 kg/m^2^ may be mistimed relative to true onset. Fourth, LTPA and MVLTPA were assessed only at baseline, implicitly assuming stability of habits, and were self-reported using different instruments across cohorts; both features can introduce misclassification and likely bias estimates toward the null, while also complicating cross-cohort comparability (partly mitigated by cohort-specific quartiles). Fifth, our MR proxy has its own constraints: the CRF-PRS instrument strength appears modest, raising weak-instrument concerns; and while we performed multiple horizontal pleiotropy checks, it cannot be completely ruled out, particularly if genetic variants influence BMI through other pathways correlated with fitness. Finally, our analyses were conducted in healthy free-living individuals of a general population in a Mediterranean country, and generalizability of our findings to other populations and ancestries should be carefully evaluated. Due to the procedure to access to BMI values, we may have also missed certain segments of the population, such as high-income individuals who primarily use private healthcare services, potentially limiting the representation of the highest socioeconomic strata.

In conclusion, higher LTPA and MVLTPA were consistently associated with more favorable lifelong BMI trajectories (slower increases until mid-adulthood, slower decreases in older adults, lower values from midlife onwards), delayed risk of obesity, and convergent support from one-sample MR analyses, supporting a causal protective role of physical activity on BMI trajectories throughout life (in both sexes and overall stronger in men). Our findings extend short/medium-term trial evidence to a life-course, real world setting and reinforce physical activity promotion as a scalable strategy for obesity prevention in primary care. Future work should validate these patterns in larger cohorts with repeated BMI measures, as well as in other ancestries and settings, incorporate repeated/objective activity and body composition measures, and evaluate implementation and cost-effectiveness of long-term activity-promotion interventions.

## Data Availability

All data produced in the present study are available upon reasonable request to the authors. Requests should be addressed to the CORDELIA Principal Investigator.

## ACKNOWLEDGMENTS

The CORDELIA team thanks: (1) all participants from the individual cohorts; (2) the health personnel and fieldwork coordinators who enabled collection of data and biological samples; (3) Marta Cabañero for building the study database; (4) Esmeralda Gómez for project management; (5) the Hospital del Mar Research Institute laboratory team (Daniel Muñoz-Aguayo, Gemma Blanchart, and Sònia Gaixas) for DNA extraction and quality control; and (6) the teams at the Centro Nacional de Genotipado and the Centro Singular de Investigación en Medicina Molecular y Enfermedades Crónicas for genotyping and support with genotyped data quality control. CIBER de Enfermedades Cardiovasculares (CIBERCV) and CIBER de Enfermedades Raras (CIBERER) are initiatives of Instituto de Salud Carlos III (Madrid, Spain) and are financed by the European Regional Development Fund.

## AUTHOR CONTRIBUTIONS

Conceptualization and Methodology: Á.H. Project administration: Á.H. Formal analysis: J.H.-R. Visualization: J.H.-R. Supervision: Á.H. Writing-Original draft preparation: J.H.-R. Writing-Review and editing: A.C.-V., R.E., E.F., M.B.L., P.T.-M., A.J.-N., J.M.V., V.M.L.-L., T.S.-F., S.C., R.C., J.M., and Á.H. J.H.-R. and Á.H. had full access to all the data in the study and take responsibility for its integrity and the data analysis. All authors read and approved the final manuscript.

## ETHICS

This study was performed in line with the principles of the Declaration of Helsinki. Approval for the study was granted by the Ethics Committee of the Parc de Salut Mar (2024/11670/I, date: November 18, 2024). Approval for the CORDELIA Study was granted by the Ethics Committee of the Parc de Salut Mar (2023/10785/I, date: March 29, 2023). All cohorts obtained written informed consent by their participants and were approved by the corresponding ethics committees.

## DATA AVAILABILITY

Due to national data-protection legislation and ethical restrictions, and because participants did not provide explicit written consent for open data sharing, the underlying study dataset cannot be deposited in a public repository. However, access for the purposes of bona fide collaborative research may be granted to qualified investigators upon reasonable request and subject to appropriate approvals and data-sharing agreements. Requests should be addressed to the CORDELIA Principal Investigator. All code used for database construction, data management, and statistical analyses is publicly available at https://github.com/XXX.

## References

1. Zhou X-D, Chen Q-F, Yang W, Zuluaga M, Targher G, Byrne CD, et al. Burden of disease attributable to high body mass index: an analysis of data from the Global Burden of Disease Study 2021. eClinicalMedicine. 2024;76:102848. 10.1016/j.eclinm.2024.102848

2. Wu Z, Xia F, Wang W, Zhang K, Fan M, Lin R. The global burden of disease attributable to high body mass index in 204 countries and territories from 1990 to 2021 with projections to 2050: An analysis of the Global Burden of Disease Study 2021. European J of Heart Fail. 2025;27:354–65. 10.1002/ejhf.3539

3. OECD. Healthy Eating and Active Lifestyles: Best Practices in Public Health. OECD; 2022 June. 10.1787/40f65568-en

4. Donnelly JE, Blair SN, Jakicic JM, Manore MM, Rankin JW, Smith BK. Appropriate Physical Activity Intervention Strategies for Weight Loss and Prevention of Weight Regain for Adults: Medicine & Science in Sports & Exercise [Internet]. 2009 [cited 2026 Feb 4];41:459–71. 10.1249/MSS.0b013e3181949333

5. Batrakoulis A, Jamurtas AZ, Metsios GS, Perivoliotis K, Liguori G, Feito Y, et al. Comparative Efficacy of 5 Exercise Types on Cardiometabolic Health in Overweight and Obese Adults: A Systematic Review and Network Meta-Analysis of 81 Randomized Controlled Trials. Circ: Cardiovascular Quality and Outcomes [Internet]. 2022 [cited 2025 Oct 29];15. 10.1161/CIRCOUTCOMES.121.008243

6. Ghoreishy SM, Noormohammadi M, Zeraattalab-Motlagh S, Shoaibinobarian N, Hasan Rashedi M, Movahed S, et al. The Effectiveness of Nonsurgical Interventions for Weight Loss Maintenance in Adults: An Updated, GRADE-Assessed Systematic Review and Meta-Analysis of Randomized Clinical Trials. Nutrition Reviews. 2025;83:809–18. 10.1093/nutrit/nuae128

7. Men J, Wang P, Gao Q, Li Y, Zhu G, Yu Z, et al. Impact of exercise on anthropometric outcomes in children and adolescents with overweight or obesity: a systematic review and meta-analysis based on 113 randomized controlled trials worldwide. BMC Public Health. 2025;25:2400. 10.1186/s12889-025-23413-9

8. Fewell Z, Davey Smith G, Sterne JAC. The Impact of Residual and Unmeasured Confounding in Epidemiologic Studies: A Simulation Study. American Journal of Epidemiology. 2007;166:646–55. 10.1093/aje/kwm165

9. Geidl W, Schlesinger S, Mino E, Miranda L, Pfeifer K. Dose–response relationship between physical activity and mortality in adults with noncommunicable diseases: a systematic review and meta-analysis of prospective observational studies. Int J Behav Nutr Phys Act. 2020;17:109. 10.1186/s12966-020-01007-5

10. Lawlor DA, Tilling K, Davey Smith G. Triangulation in aetiological epidemiology. Int J Epidemiol [Internet]. 2017 [cited 2025 May 22];dyw314. 10.1093/ije/dyw314

11. Artime E, Spaepen E, Zimner-Rapuch S, Lampropoulou A, Adam A, Lin X, et al. Epidemiology Landscape and Impact of Overweight and Obesity in Adults: Multi-country Results from the IMPACT-O Study. Adv Ther [Internet]. 2025 [cited 2026 Feb 5];42:5148–63. 10.1007/s12325-025-03333-1

12. Davey Smith G, Hemani G. Mendelian randomization: genetic anchors for causal inference in epidemiological studies. Human Molecular Genetics. 2014;23:R89–98. 10.1093/hmg/ddu328

13. Burgess S, Davey Smith G, Davies NM, Dudbridge F, Gill D, Glymour MM, et al. Guidelines for performing Mendelian randomization investigations: update for summer 2023. Wellcome Open Res. 2023;4:186. 10.12688/wellcomeopenres.15555.3

14. Hanscombe KB, Persyn E, Traylor M, Glanville KP, Hamer M, Coleman JRI, et al. The genetic case for cardiorespiratory fitness as a clinical vital sign and the routine prescription of physical activity in healthcare. Genome Med. 2021;13:180. 10.1186/s13073-021-00994-9

15. Schröder H, Subirana I, Elosua R, Camps-Vilaró A, Tizón-Marcos H, Fitó M, et al. Measuring Cardiorespiratory Fitness without Exercise Testing: The Development and Validation of a New Tool for Spanish Adults. JCM. 2024;13:2210. 10.3390/jcm13082210

16. Hernáez Á, Camps-Vilaró A, Polo-Alonso S, Subirana I, Ramos R, De Cid R, et al. Cohort profile: the CORDELIA study (Collaborative cOhorts Reassembled Data to study mEchanisms and Longterm Incidence of chronic diseAses). Eur J Epidemiol [Internet]. 2025 [cited 2025 June 11]; 10.1007/s10654-025-01229-6

17. Elosua R, Marrugat J, Molina L, Pons S, Pujol E. Validation of the Minnesota Leisure Time Physical Activity Questionnaire in Spanish Men. American Journal of Epidemiology. 1994;139:1197–209.

18. Elosua R, Garcia M, Aguilar A, Molina L, Covas M-I, Marrugat J. Validation of the Minnesota Leisure Time Physical Activity Questionnaire in Spanish Women: Medicine & Science in Sports & Exercise [Internet]. 2000 [cited 2025 Mar 5];32:1431–7. 10.1097/00005768-200008000-00011

19. Rial-Vázquez J, Pérez-Ríos M, Santiago-Pérez MI, Ruano-Ravina A. Versión reducida del Minnesota Leisure Time Physical Activity Questionnaire para población general: MLTPAQ 9 + 2. Gaceta Sanitaria [Internet]. 2023 [cited 2025 June 11];37:102309. 10.1016/j.gaceta.2023.102309

20. Maddison R, Ni Mhurchu C, Jiang Y, Vander Hoorn S, Rodgers A, Lawes CM, et al. International Physical Activity Questionnaire (IPAQ) and New Zealand Physical Activity Questionnaire (NZPAQ): a doubly labelled water validation. Int J Behav Nutr Phys Act. 2007;4:62. 10.1186/1479-5868-4-62

21. Ge T, Chen C-Y, Ni Y, Feng Y-CA, Smoller JW. Polygenic prediction via Bayesian regression and continuous shrinkage priors. Nat Commun. 2019;10:1776. 10.1038/s41467-019-09718-5

22. Agència de Qualitat i Avaluació Sanitàries de Catalunya (AQuAS)-Generalitat de Catalunya. Programa d’analítica de dades per a la recerca i la innovació en salut (PADRIS) 2022. [Internet]. https://aquas.gencat.cat/ca/detall/article/padris

23. Instituto Nacional de Estadística (INE). Atlas de Distribución de Renta de los Hogares (2015-2023) [Internet]. Madrid, España; 2024. https://www.ine.es/experimental/atlas/experimental_atlas.htm

24. Elhakeem A, Hughes RA, Tilling K, Cousminer DL, Jackowski SA, Cole TJ, et al. Using linear and natural cubic splines, SITAR, and latent trajectory models to characterise nonlinear longitudinal growth trajectories in cohort studies. BMC Med Res Methodol [Internet]. 2022 [cited 2025 June 10];22:68. 10.1186/s12874-022-01542-8

25. Pérez-Vega K-A, Lassale C, Zomeño M-D, Castañer O, Salas-Salvadó J, Basterra-Gortari FJ, et al. Breakfast energy intake and dietary quality and trajectories of cardiometabolic risk factors in older adults. The Journal of nutrition, health and aging. 2024;28:100406. 10.1016/j.jnha.2024.100406

26. Zhang X, Molsberry SA, Schwarzschild MA, Ascherio A, Gao X. Association of Diet and Physical Activity With All-Cause Mortality Among Adults With Parkinson Disease. JAMA Netw Open. 2022;5:e2227738. 10.1001/jamanetworkopen.2022.27738

27. Price AL, Patterson NJ, Plenge RM, Weinblatt ME, Shadick NA, Reich D. Principal components analysis corrects for stratification in genome-wide association studies. Nat Genet [Internet]. 2006 [cited 2026 Feb 5];38:904–9. 10.1038/ng1847

28. Lawlor DA, Tilling K, Davey Smith G. Triangulation in aetiological epidemiology. Int J Epidemiol. 2017;dyw314. 10.1093/ije/dyw314

29. Von Hinke S, Davey Smith G, Lawlor DA, Propper C, Windmeijer F. Genetic markers as instrumental variables. Journal of Health Economics. 2016;45:131–48. 10.1016/j.jhealeco.2015.10.007

30. Lipsitch M, Tchetgen Tchetgen E, Cohen T. Negative Controls: A Tool for Detecting Confounding and Bias in Observational Studies. Epidemiology. 2010;21:383–8. 10.1097/EDE.0b013e3181d61eeb

31. Hemani G, Tilling K, Davey Smith G. Orienting the causal relationship between imprecisely measured traits using GWAS summary data. Li J, editor. PLoS Genet. 2017;13:e1007081. 10.1371/journal.pgen.1007081

32. Sanderson E, Glymour MM, Holmes MV, Kang H, Morrison J, Munafò MR, et al. Mendelian randomization. Nat Rev Methods Primers [Internet]. 2022 [cited 2025 June 10];2:6. 10.1038/s43586-021-00092-5

33. Bellicha A, Van Baak MA, Battista F, Beaulieu K, Blundell JE, Busetto L, et al. Effect of exercise training on weight loss, body composition changes, and weight maintenance in adults with overweight or obesity: An overview of 12 systematic reviews and 149 studies. Obesity Reviews [Internet]. 2021 [cited 2025 Sept 4];22:e13256. 10.1111/obr.1325634.

34. Cleven L, Krell-Roesch J, Nigg CR, Woll A. The association between physical activity with incident obesity, coronary heart disease, diabetes and hypertension in adults: a systematic review of longitudinal studies published after 2012. BMC Public Health [Internet]. 2020 [cited 2025 June 10];20:726. 10.1186/s12889-020-08715-4

35. Langsetmo L, Hitchcock CL, Kingwell EJ, Davison KS, Berger C, Forsmo S, et al. Physical activity, body mass index and bone mineral density—associations in a prospective population-based cohort of women and men: The Canadian Multicentre Osteoporosis Study (CaMos). Bone [Internet]. 2012 [cited 2025 Aug 7];50:401–8. 10.1016/j.bone.2011.11.009

36. Duncan GE, Cash SW, Horn EE, Turkheimer E. Quasi-causal associations of physical activity and neighborhood walkability with body mass index: A twin study. Preventive Medicine [Internet]. 2015 [cited 2025 Oct 3];70:90–5. 10.1016/j.ypmed.2014.11.024

37. Hill JO, Wyatt HR, Peters JC. Energy Balance and Obesity. Circulation [Internet]. 2012 [cited 2026 Feb 20];126:126–32. 10.1161/CIRCULATIONAHA.111.087213

38. Yerrakalva D, Hajna S, Khaw K-T, Griffin SJ, Brage S. Prospective associations between changes in physical activity and sedentary time and subsequent lean muscle mass in older English adults: the EPIC-Norfolk cohort study. Int J Behav Nutr Phys Act [Internet]. 2024 [cited 2026 Feb 20];21:10. 10.1186/s12966-023-01547-6

39. Herzig S, Shaw RJ. AMPK: guardian of metabolism and mitochondrial homeostasis. Nat Rev Mol Cell Biol [Internet]. 2018 [cited 2026 Feb 20];19:121–35. 10.1038/nrm.2017.95

40. Strain T, Dempsey PC, Wijndaele K, Sharp SJ, Kerrison N, Gonzales TI, et al. Quantifying the Relationship Between Physical Activity Energy Expenditure and Incident Type 2 Diabetes: A Prospective Cohort Study of Device-Measured Activity in 90,096 Adults. Diabetes Care [Internet]. 2023 [cited 2026 Feb 20];46:1145–55. 10.2337/dc22-1467

41. Caudwell P, Gibbons C, Finlayson G, Näslund E, Blundell J. Exercise and Weight Loss: No Sex Differences in Body Weight Response to Exercise. Exercise and Sport Sciences Reviews [Internet]. 2014 [cited 2026 Feb 20];42:92–101. 10.1249/JES.0000000000000019

42. Christensen P, Meinert Larsen T, Westerterp-Plantenga M, Macdonald I, Martinez JA, Handjiev S, et al. Men and women respond differently to rapid weight loss: Metabolic outcomes of a multi-centre intervention study after a low-energy diet in 2500 overweight, individuals with pre-diabetes (PREVIEW). Diabetes Obesity Metabolism [Internet]. 2018 [cited 2026 Feb 20];20:2840–51. 10.1111/dom.13466

43. Sternfeld B, Bhat AK, Wang H, Sharp T, Quesenberry CP. Menopause, Physical Activity, and Body Composition/Fat Distribution in Midlife Women. Medicine & Science in Sports & Exercise [Internet]. 2005 [cited 2026 Feb 20];37:1195–202. 10.1249/01.mss.0000170083.41186.b1

44. Slootmaker SM, Schuit AJ, Chinapaw MJ, Seidell JC, Van Mechelen W. Disagreement in physical activity assessed by accelerometer and self-report in subgroups of age, gender, education and weight status. Int J Behav Nutr Phys Act [Internet]. 2009 [cited 2026 Feb 20];6:17. 10.1186/1479-5868-6-17

45. The InterAct Consortium. Validity of a short questionnaire to assess physical activity in 10 European countries. Eur J Epidemiol [Internet]. 2012 [cited 2026 Feb 20];27:15–25. 10.1007/s10654-011-9625-y

