## Supplemental material for "Leisure-time physical activity on lifelong trajectories of body mass index and obesity risk throughout life: multivariable regression and Mendelian randomization analyses using real-world data from the CORDELIA-Catalunya Study"

**SUPPLEMENTARY MATERIALS**

**Supplementary Table 1.** Number of BMI measurements per individual distributed by 10-year age group (30–90) and percentage of total values

| **Age (10-year interval)** | **Count** |
| --- | --- |
| [30,40) | 1,686 (4.7%) |
| [40,50) | 5,688 (15.7%) |
| [50,60) | 10,132 (28.0%) |
| [60,70) | 9,793 (27.1%) |
| [70,80) | 6,352 (17.6%) |
| [80,90) | 2,506 (6.9%) |

**Supplementary Table 2.** Baseline characteristics in men and women separately

|  | **Men**  *n* = 7,090 | **Women**  *n* = 7,903 |
| --- | --- | --- |
| Age at baseline, years (mean ± SD) | 53.0 ± 11.2 | 53.9 ± 10.9 |
| Baseline BMI, kg/m^2^ (mean ± SD) | 27.8 ± 4.03 | 27.3 ± 5.06 |
| Baseline prevalence of diabetes (*n*, %) | 556 (7.84%) | 438 (5.54%) |
| Baseline prevalence of hypertension (*n*, %) | 2,988 (42.1%) | 3,275 (41.4%) |
| Baseline prevalence of hypercholesterolemia (*n*, %) | 2,992 (42.2%) | 3,277 (41.5%) |
| Smoking status: |  |  |
| Current smoker (*n*, %) | 2,310 (32.6%) | 1,542 (19.5%) |
| Never smoked (*n*, %) | 2,002 (28.2%) | 5,134 (65.0%) |
| Former smoker (*n*, %) | 2,778 (39.2%) | 1,227 (15.5%) |
| Socioeconomic status, €/year (median, 1^st^-3^rd^ quartile) | 13,161 (11933; 14755) | 13,161 (11949; 14910) |

**Supplementary Table 3.** Baseline characteristics by cohort

|  | **REGICOR + ACRISC** *n* = 7,558 | **ARTPER** *n* = 1,270 | **ILERVAS** *n* = 6,165 |
| --- | --- | --- | --- |
| Age at baseline, years (mean ± SD) | 54.7 ± 12.9 | 62.5 ± 7.78 | 50.1 ± 7.06 |
| Baseline BMI, kg/m^2^ (mean ± SD) | 27.1 ± 4.61 | 29.0 ± 4.48 | 27.8 ± 4.56 |
| Baseline prevalence of diabetes (*n*, %) | 797 (10.5%) | 197 (15.5%) | 0 (0.00%) |
| Baseline prevalence of hypertension (*n*, %) | 2,250 (29.8%) | 573 (45.1%) | 2,518 (40.8%) |
| Baseline prevalence of hypercholesterolemia (*n*, %) | 2,288 (30.3%) | 617 (48.6%) | 3,358 (54.5%) |
| Smoking status: |  |  |  |
| Current smoker (*n*, %) | 1,810 (23.9%) | 212 (16.7%) | 1,830 (29.7%) |
| Never smoked (*n*, %) | 4,010 (53.1%) | 694 (54.6%) | 2,432 (39.4%) |
| Former smoker (*n*, %) | 1,738 (23.0%) | 364 (28.7%) | 1,903 (30.9%) |
| Socioeconomic status, €/year (median, 1^st^-3^rd^ quartile) | 14,604  (13,364; 14,910) | 11,933  (10,748; 14,172) | 12,108  (11,688; 12,916) |

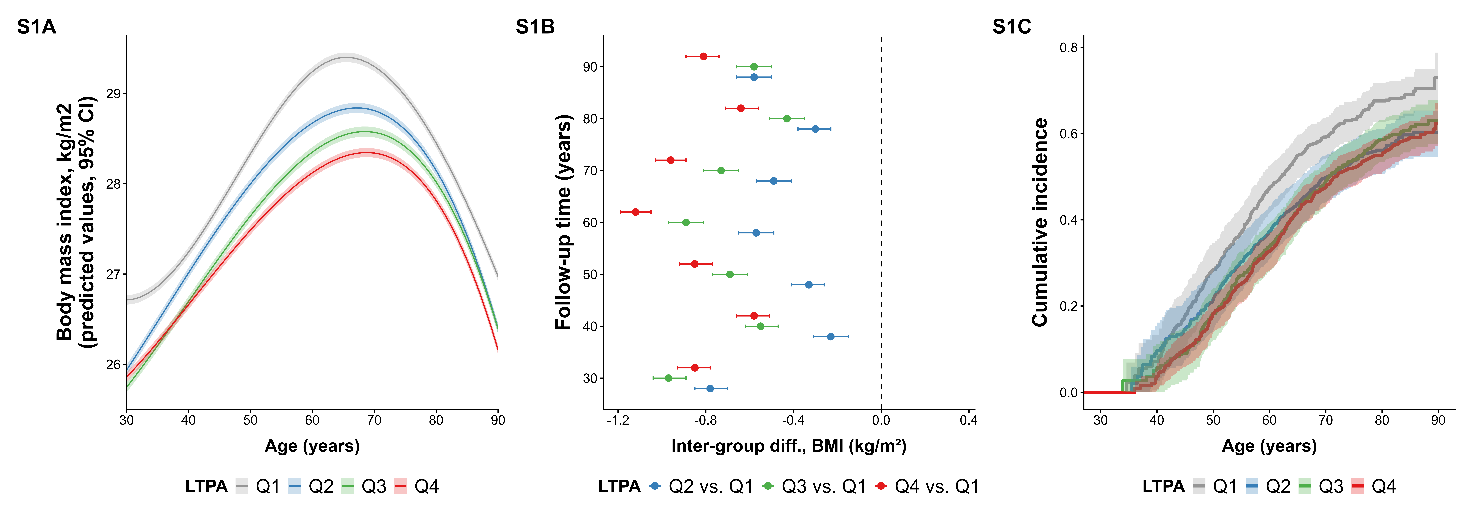
**Supplemental Figure 1.** BMI trajectories and incident obesity by cohort-specific quartiles of LTPA only in men (Q1 in grey, Q2 in blue, Q3 in green, Q4 in red). A. BMI trajectories per quartiles. B. Inter-group differences in predicted mean BMI values between ages 30-90. C. Weighted Kaplan-Meier curves for first obesity onset.

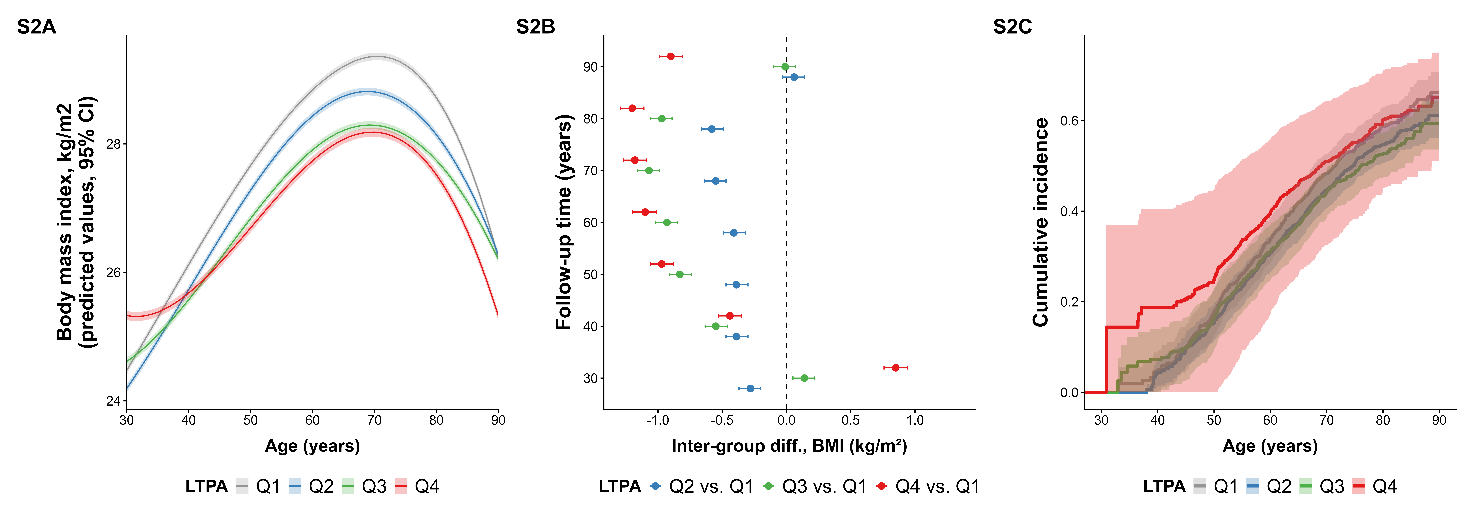
**Supplemental Figure 2.** BMI trajectories and incident obesity by cohort-specific quartiles of LTPA only in women (Q1 in grey, Q2 in blue, Q3 in green, Q4 in red). A. BMI trajectories per quartiles. B. Inter-group differences in predicted mean BMI values between ages 30-90. C. Weighted Kaplan-Meier curves for first obesity onset.

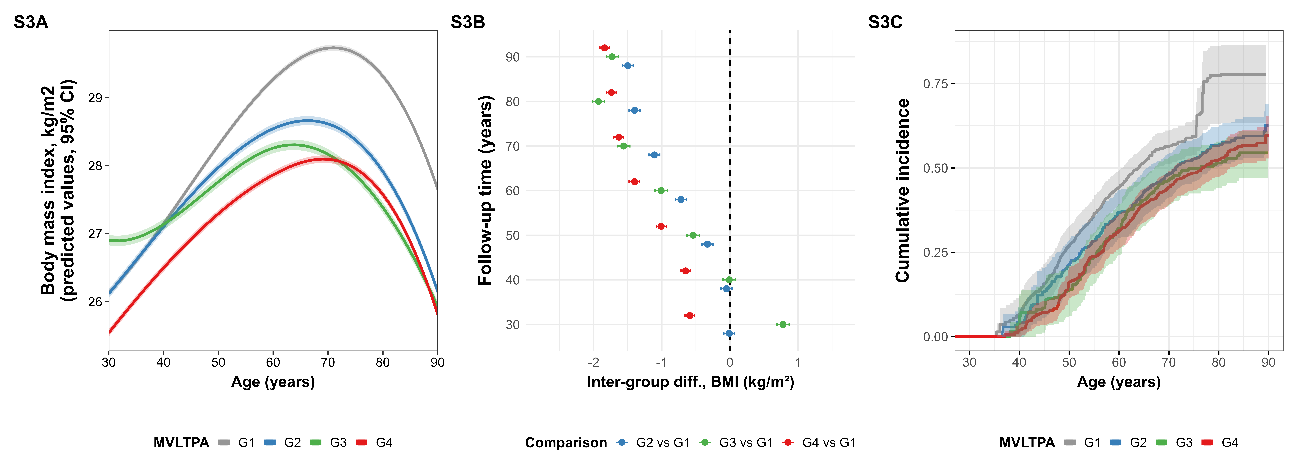
**Supplemental Figure 3.** BMI trajectories and incident obesity by MVLTPA levels in REGICOR-ACRISC only in men (0 METs-min/day in grey, >0 to <100 METs-min/day in blue, ≥100 to <200 METs-min/day in green, ≥200 METs-min/day in red). A. BMI trajectories per quartiles. B. Inter-group differences in predicted mean BMI values between ages 30-90. C. Weighted Kaplan-Meier curves for first obesity onset.

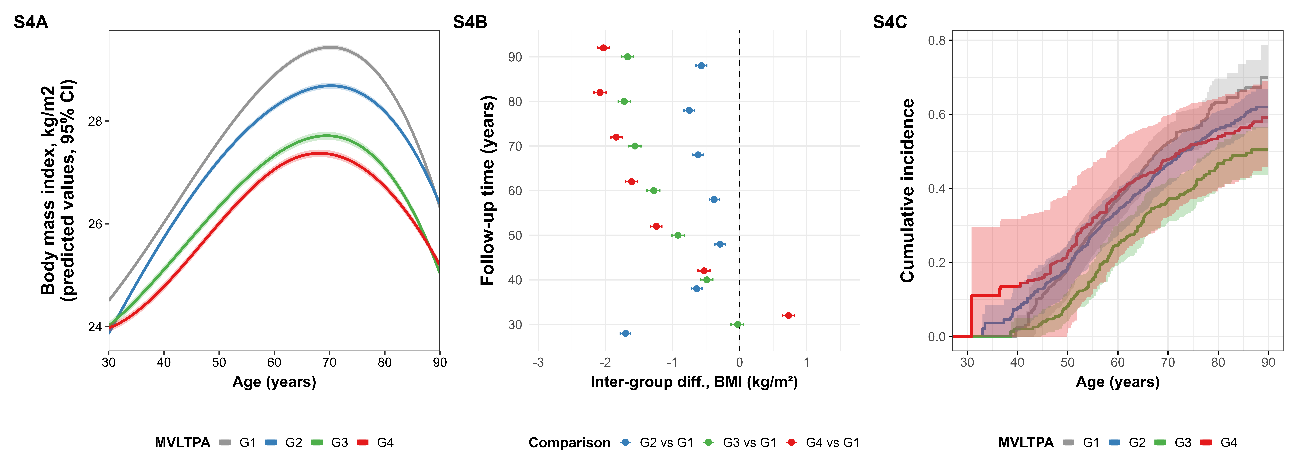
**Supplemental Figure 4.** BMI trajectories and incident obesity by MVLTPA levels in REGICOR-ACRISC only in women (0 METs-min/day in grey, >0 to <100 METs-min/day in blue, ≥100 to <200 METs-min/day in green, ≥200 METs-min/day in red). A. BMI trajectories per quartiles. B. Inter-group differences in predicted mean BMI values between ages 30-90. C. Weighted Kaplan-Meier curves for first obesity onset.

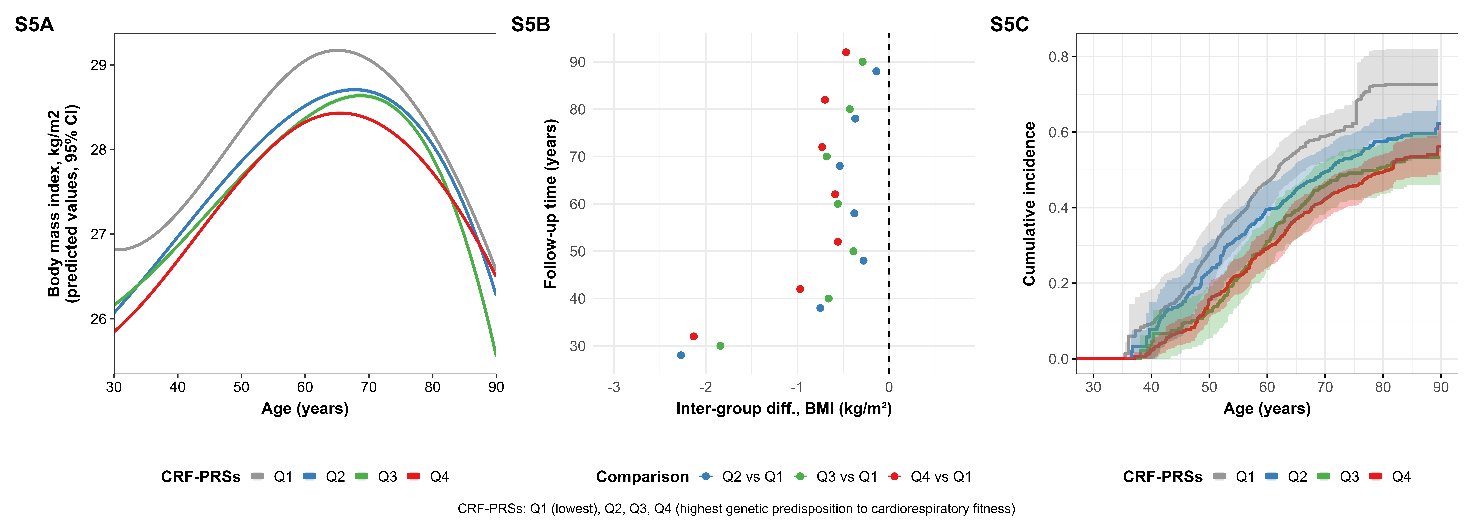
**Supplemental Figure 5.** BMI trajectories and incident obesity by genetically determined CRF levels only in men (Q1 in grey, Q2 in blue, Q3 in green, Q4 in red). A. BMI trajectories per quartiles. B. Inter-group differences in predicted mean BMI values between ages 30-90. C. Weighted Kaplan-Meier curves for first obesity onset.

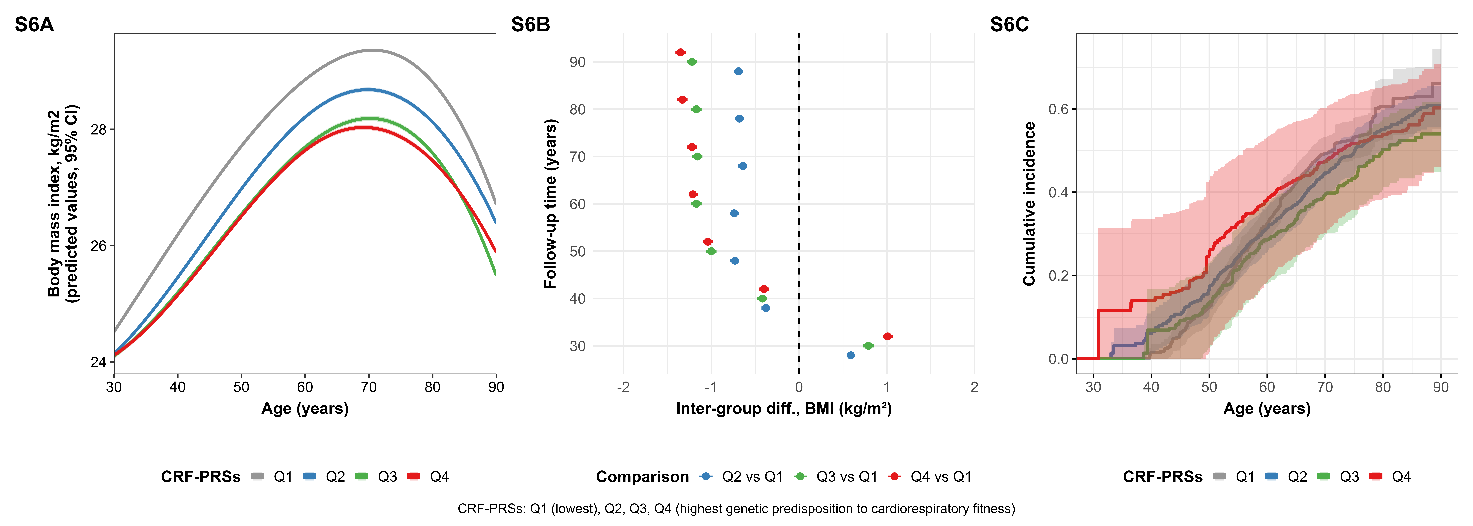
**Supplemental Figure 6.** BMI trajectories and incident obesity by genetically determined CRF levels only in women (Q1 in grey, Q2 in blue, Q3 in green, Q4 in red). A. BMI trajectories per quartiles. B. Inter-group differences in predicted mean BMI values between ages 30-90. C. Weighted Kaplan-Meier curves for first obesity onset.
